# A framework for guiding integrated disease control measures through multipathogen surveillance

**DOI:** 10.1101/2025.11.02.25339351

**Authors:** Samantha J. Bents, Pearl Anne Ante-Testard, Huiyu Hu, Francois Rerolle, Everlyn Kamau, Sammy M. Njenga, Henry Kanyi, Richelle W. Kihoro, Henry Emisiko, Mahbubur Rahman, Sania Ashraf, Sarker Masud Parvez, Shahjahan Ali, Md. Ziaur Rahman, Md. Rana Miah, Mostofa Shakil, Abdul Kashan Shoab, Rashidul Haque, E. Brook Goodhew, Chabier Coleman, Jessica A. Grembi, Andrew N. Mertens, Christine P. Stewart, Jeffrey W. Priest, Patrick J. Lammie, Audrie Lin, Jade Benjamin-Chung, Ayse Ercumen, Amy J. Pickering, Clair Null, John M. Colford, Stephen P. Luby, Nathan C. Lo, Benjamin F. Arnold

## Abstract

Global health programs have traditionally focused on single diseases. There is potential for synergy through integrated intervention delivery, particularly in areas with overlapping geographic disease burden, but there is limited methodology developed to guide integration. Here, we propose a framework to translate multipathogen surveillance data into summary indices that can guide integrated program delivery to target multiple pathogens at once. We considered several measures of diversity to quantify multipathogen infection or undervaccination across three large-scale surveys: subregions in Bangladesh (90 clusters, n = 7,187) and Kenya (89 clusters, n = 9,077), and a national survey in Cambodia (100 clusters, n = 2,150). We found that strategies guided by a measure of diversity, Rao’s quadratic index, consistently identified more efficient integrated strategies than other multipathogen indices across a range of intervention combinations. We then identified case studies in each setting that used immunization as the backbone for integration and found that incorporating information from co-occurring pathogens led to only modest reductions in efficiency for vaccine-preventable diseases, while affording potentially large gains for other pathogens. Compared with a strategy that prioritized a single vaccine-preventable disease, prioritization using a multipathogen index required targeting 4% (95% CI: 0-12%) fewer spatial clusters in Bangladesh to target four additional pathogens, 10% fewer clusters in Kenya (95% CI: 3-17%), and 11% (95% CI: 5-22%) fewer clusters in Cambodia to reach 80% of all potential intervention beneficiaries across diseases. In all settings, we observed geographic clustering of multiple pathogens, with moderate to high spatial autocorrelation in Bangladesh (Moran’s I = 0.58), Kenya (I = 0.56), and Cambodia (I = 0.26), indicating potential for more efficient, spatially concentrated integrated interventions. We also found that a multipathogen index was significantly associated with a multidimensional poverty index in Bangladesh (Spearman’s ρ = 0.25), Kenya (ρ = 0.51), and Cambodia (ρ = 0.59), suggesting that multipathogen-guided strategies may effectively target impoverished populations with the highest public health need. The framework provides a systematic approach to guide integrated program delivery, offering potential for greater efficiency in disease control.

## Introduction

Historically, disease monitoring and mitigation efforts have focused on a single pathogen or concentrated only on smaller subsets of related pathogens. However, advancements in multipathogen surveillance and diagnostics have made it more feasible to implement broader monitoring and mitigation strategies in complex global health settings^1–5^. The simultaneous measurement of multiple pathogens can reveal populations with high geographically overlapping exposure to several infectious diseases, highlighting populations experiencing multilayered health disparities^6,7^. In settings where multiple pathogens overlap, integrated program delivery could enable more efficient control than independent programs with a single-pathogen focus, yet limited work has been done in methodology to guide integrated intervention delivery.

The World Health Organization identified the integration of disease interventions as a central pillar of its 2021-2030 roadmap to prevent, control, and eliminate 20 diseases and broader disease groups^8^. The roadmap outlined the need for coordination across vector control programs, immunization campaigns, water, sanitation, and hygiene (WASH) interventions, and other disease-specific programs. Coordinated interventions could more efficiently target the complex multipathogen disease burden that communities experience, while also reducing the logistical challenges and financial costs associated with delivering multiple health interventions independently. In light of the disruptions to global health programs caused by the COVID-19 pandemic and the more current restructuring of global health financing, the need for more efficient, cross-cutting interventions is a particularly urgent problem^9,10^.

Assessing the potential for integrated delivery requires condensing complex multipathogen data into actionable interventions. Statistical measures of diversity, which quantify the composition of communities by evaluating the distinct entities present and their distributions, offer concise ways to condense multidimensional data into single quantities that represent the complex diversity of the communities they measure. Accordingly, statistical measures of diversity are often used to translate complex information into more interpretable metrics that may be better suited to inform actionable policy^11^. For example, such metrics have been rigorously applied in ecological systems and conservation planning to translate rich information on species’ distributions into single indices representing the biodiversity present in a given location for conservation monitoring and prioritization^12^. In recent years, diversity metrics have also gained traction in epidemiological settings, including to quantify the intensity of disease outbreaks over time^13,14^, assess genetic and microbiome diversities^15^, guide surveillance efforts^16^, and quantify the uneven distribution of disease within populations^17^.

Integrated disease control should be advantageous in settings where individual disease distributions overlap geographically. Spatial co-occurrence of diseases can arise for various reasons. Vector-borne diseases, such as malaria, dengue, and chikungunya, often exhibit overlapping endemicity, driven in part by similar environmental conditions that support vector viability^18^. Populations living in under-resourced settings also face elevated risk for neglected tropical diseases, particularly parasitic pathogens such as soil-transmitted helminths, largely due to limited access to improved sanitation and health services^19^. Such deprivation is often spatially concentrated as a result of institutional, geographic, environmental, and structural factors^20^. Demographic and behavioral factors, such as high-contact behaviors or shared population mobility patterns, can also partially explain where infectious diseases aggregate^21,22^.

Growing investment in large-scale, population-based serosurveillance to monitor priority pathogens provides an opportunity to detect and intervene on spatially overlapping infectious diseases^23–29^. Here, we developed a framework for synthesizing high-dimensional multiplex serological surveillance data into actionable opportunities for more efficient integrated disease control. Our analysis had three objectives: first, to evaluate several candidate diversity metrics, hereafter referred to as multipathogen indices, and their ability to identify efficient integrated strategies across diverse epidemiological settings. We hypothesized that multipathogen indices may be useful tools for condensing multipathogen surveillance data to identify geographic areas with overlapping intervention needs, where integrated interventions should provide efficient gains over traditional single-pathogen targeting. Second, using the best-performing multipathogen index, we developed a framework for guiding integrated multipathogen interventions that considered realistic platform implementation, efficiency, and spatial practicality. We anticipated that we could identify integrated interventions that could be efficiently administered at spatial scales relevant to public health, given shared risk factors across infectious diseases. Lastly, we characterized shared environmental and household risk factors of populations residing in settings with overlapping intervention needs. We hypothesized that areas of high overlapping intervention need would be distinguished by identifiable risk factors, such as housing conditions and access to improved water and sanitation, which may underpin infectious disease risk and population well-being more broadly. To evaluate our framework, we used large-scale surveys tested with multiplex assays and stool-based microscopy in three settings spanning diverse epidemiological contexts: subnational surveys in Bangladesh and Kenya, and a nationally representative survey in Cambodia.

## Methodology

### Data

We analyzed large-scale multipathogen surveillance data collected in three different settings: subregions in Kenya (n = 9,077 children) and Bangladesh (n = 7,187 children), and a national survey in Cambodia (n = 2,150 adult women) (**Table 1**). Across all settings, multiplex serological data were analyzed using the Luminex Bio-Plex 200 instrument (Luminex Corp., Austin, TX). Samples from the Kenya and Bangladesh studies were tested at the Kenya Medical Research Institute in Nairobi, Kenya, and samples from Cambodia were tested at the Centers for Disease Control and Prevention in Atlanta, Georgia, USA^30^. In the Bangladesh and Kenya subregions, we additionally included multipathogen stool-based microscopy data for a subset of soil-transmitted helminths, including *Ascaris lumbricoides, Trichuris trichiura*, and hookworm^31,32^.

**Table 1.** Study population information, pathogens, and associated interventions by location.

| **Setting** | **Study population sample size (n), age range, and years of collection** | **Spatial clusters** | **Pathogens that meet prior condition can be targeted by vaccines (VPD), mass drug administration (MDA), or malaria package** |
|---|---|---|---|
| Bangladesh subregion | Multiplex serology: 4,292 children <30 months, collected 2012-2016<br>Stool-based microscopy:<br>7,187 children aged 2-12 years, collected 2015-2016 | 90 | VPD:<br>1. Measles<br>2. Rubella<br>3. Diphtheria<br>4. Tetanus<br>MDA (albendazole):<br>5. *Ascaris lumbricoides*<br>6. *Trichuris trichiura*<br>7. Hookworm |
| Kenya subregion | Multiplex serology: 5,154 children <30 months, collected 2014-2016<br>Stool-based microscopy: 9,077 children aged 2-15 years, collected 2015-2016 | 89 | VPD:<br>1. Rubella<br>2. Measles<br>MDA (e.g., albendazole or praziquantel):<br>3. *Ascaris lumbricoides*<br>4. *Taenia solium*<br>5. Schistosomiasis<br>Malaria:<br>6. *Plasmodium falciparum*<br>7. *Plasmodium malariae*<br>8. *Plasmodium ovale* |
| Cambodia | Multiplex serology: 2,150 women aged 15-39 years, collected 2012 | 100 | VPD:<br>1. Tetanus<br>MDA (ivermectin):<br>2. *Strongyloides stercoralis*<br>3. Lymphatic filariasis<br>Malaria:<br>4. *Plasmodium falciparum*<br>5. *Plasmodium vivax* |

### Seropositivity cutoffs

For each serological dataset, individual seropositivity was determined using binary cutoff values. In Cambodia, seropositivity for each pathogen was defined using cutoff values derived from negative control human sera, sera from parasitologically-confirmed cases, and international standard tetanus sera^30^. In the Bangladesh and Kenya subregions, seropositivity cutoffs were derived from negative controls with no history of foreign travel for malaria and lymphatic filariasis antigens, WHO international standards for seroprotection against VPDs, and receiver operating characteristic (ROC) curve analysis using known positive and negative reference sera for the remaining antigens. Individuals were considered seropositive if their antibody level exceeded the established seropositivity cutoff for at least one antigen for a given pathogen, except lymphatic filariasis where seropositivity to both Bm14 and Wb123 antigens was required^30,33^.

### Defining intervention need

From each individual’s serostatus, we assigned a binary indicator reflecting whether that individual would benefit from an intervention treating each of the pathogens considered, where beneficiaries were either seronegative to vaccine-specific antigens (suggesting missing or incomplete protection from vaccination), or seropositive to other pathogens, indicating likely recent exposure. For the serological datasets in the Bangladesh and Kenya subregions, the young age of study participants ensured that serostatus was indicative of recent exposure. In Cambodia, the older age range of the population reduces the certainty that IgG levels reflect recent or past exposure; this analysis considered all antigen markers available. Estimation of pathogen-specific intervention needs are provided in **Supplementary Figure S1.**

From the full set of pathogens tested across diagnostic platforms, the analysis focused on pathogens meeting two conditions: 1) pathogens for which ≥5% of the population could benefit from its associated intervention, and 2) pathogens that could be addressed with delivery of an intervention, which were grouped broadly into three categories including vaccine-preventable diseases (VPD), mass drug administrations (MDA), and malaria packages (e.g., vector control, MDA, or vaccination). More detailed recommended guidelines for the control of each study pathogen are provided in **Supplementary Table S1,** and information on the full number of pathogens tested in each setting is provided in **Supplementary Table S2.**

### Comparing candidate multipathogen indices and their ability to identify efficient strategies

We compared the efficiency of integrated strategies combining single-pathogen interventions when guided by four different diversity metrics, hereafter referred to as multipathogen indices: species richness *R*(*s*), corresponding to the number of intervenable pathogens present in a given spatial cluster, Shannon diversity index *H*(*s*), Gini-Simpson index *D*(*s*), and Rao’s quadratic index *Q*(*s*). At each spatial cluster *s,* these multipathogen indices were calculated using the following equations:

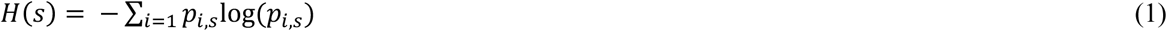

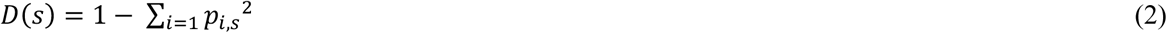

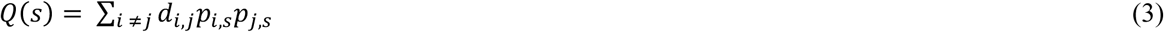

Shannon and Gini-Simpson indices require a discrete probability distribution, so when computing *H*(*s*) and *D*(*s*) *p* represents pathogen *i*’s relative contribution to the total intervention need in cluster *s.* As a result, H(*s*) and D(*s*) describe the composition of intervenable pathogens within a cluster, but not the overall magnitude of intervention need, whereby H(*s*) captures richness and is sensitive to rare pathogens, and D(*s*) represents the probability that two randomly selected individuals could benefit from interventions targeting two different pathogens. In contrast, for Rao’s quadratic index, Q(*s*) was calculated from unnormalized spatial cluster-level prevalences and can be interpreted as the sum of expected co-occurrence probabilities under independence. Of note, if Q(*s*) were computed from the normalized proportions pᵢ(*s*), with *d_i,j_*= 1 for all i ≠ j, it would be equivalent to D(*s*) and yield identical cluster rankings. Here, we computed Q(*s*) from unnormalized prevalences so that it jointly reflects the composition and magnitude of intervention need. For *Q*(*s*), we specified that *d_i_*_,j_ = 1 for all pathogen pairs that could be included in a hypothetical integrated intervention and *d_i_*_,j_ = 0 otherwise. In the main analysis, we evaluated every possible combination of integrated interventions in each setting, spanning the full range of combinations available across intervenable pathogens.

In a resource-limited context, we assumed that an intervention strategy would prioritize spatial clusters with the largest number of intervention beneficiaries. Accordingly, we ranked spatial clusters in descending order of *R*(*s*), *H*(*s*), *D*(*s*), *and Q*(*s*) and calculated the cumulative percentage of overall beneficiaries reached as additional clusters are sequentially targeted when guided by each multipathogen index. To explicitly quantify the efficiency of strategies guided by these indices, we calculated the minimum number of spatial clusters needed to be included in a hypothetical integrated strategy to target at least 80% of all individuals in the population who would benefit from an intervention, whereby a more efficient strategy should be able to target more beneficiaries by visiting fewer locations. A benchmark of 80% was chosen as it corresponds to the WHO Reaching Every District target for immunization coverage^34,35^. We considered a range of other benchmarks in supplementary analyses (**Supplementary Figure S2**). In the main analysis, efficiency was defined only by the method’s ability to identify the optimal spatial clusters for integrated delivery. In supplementary analyses, we assessed a more complex approach to quantify Q(s) that additionally considered operational feasibility, where *d_i_*_,j_ = 1 only when two pathogens shared an intervention exactly (e.g., vector control targeting both malaria and lymphatic filariasis, or ivermectin MDA targeting both *Strongyloides stercoralis* and lymphatic filariasis), providing insights into strategies that would be efficient spatially and also shared synergistic interventions (**Supplementary Methods 1**).

We investigated the potential efficiency of integrating at least three pathogens in each integrated intervention, meaning that the number of total combinations *c* assessed in each location can be written as:

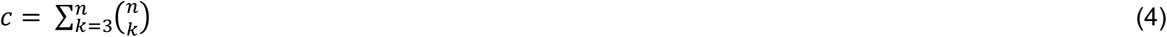

Where *n* = the total number of pathogens considered for integration, corresponding to *n* = 7 pathogens in the Bangladesh subregion (measles, rubella, diphtheria, tetanus, *Ascaris lumbricoides, Trichuris trichiura*, hookworm) *n* = 8 pathogens in the Kenya subregion (measles, rubella, *Ascaris lumbricoides, Taenia solium*, schistosomiasis, *Plasmodium falciparum*, *Plasmodium malariae*, *Plasmodium ovale*), and *n* = 5 pathogens in Cambodia (tetanus, *Strongyloides stercoralis*, lymphatic filariasis, *Plasmodium falciparum*, *Plasmodium vivax)*, and *k* is the number of pathogens included in each combination. The objective of this analysis was to evaluate which multipathogen indices could identify efficient multipathogen targeting opportunities over a wide range of possible intervention combinations.

### Framework for identifying opportunities for integrated strategies

After evaluating the best-performing multipathogen index to guide spatial prioritization, we developed a generalized framework for condensing multipathogen surveillance data into actionable opportunities for more efficient integrated disease control (**Figure 1**). The framework operates on the assumption that a multipathogen survey has been previously collected with geographic information available at the unit of data collection (**Figure 1**, Step 1), and that pathogens with potential for integration have been identified (**Figure 1**, Step 2). In this analysis, we specified that pathogens had potential for integration if they could be treated with current, large-scale public health interventions and if ≥5% of the population could benefit from the associated intervention (i.e., vaccination, malaria control, or MDA, defined in **Table 1**).

**Figure 1.**
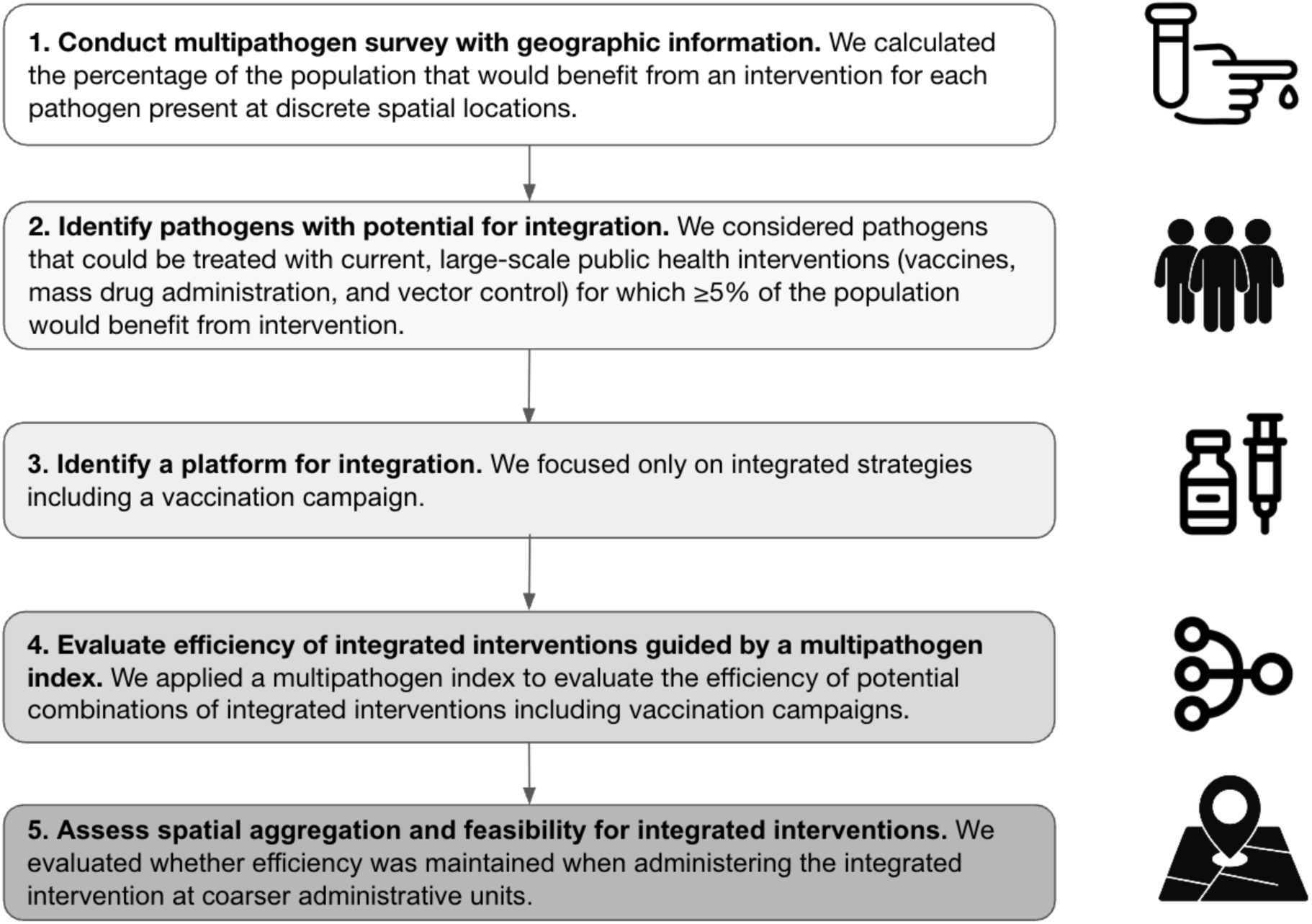
A proposed framework to identify opportunities for integrated interventions from multipathogen surveillance data. The framework requires that a multipathogen survey has been conducted with spatial information. It then considers integration potential, efficiency, and spatial operationality to identify pragmatic opportunities for integration.

Integrated programs will likely be most feasible if they build from existing, single-intervention platforms (**Figure 1**, Step 3). We used vaccination campaigns as an example because supplemental vaccine campaigns are well-developed and widespread in resource-limited settings^36,37^. Accordingly, we only considered integrated strategies that appended additional interventions such as malaria control and MDA to existing vaccine campaigns, corresponding to childhood measles immunization in Bangladesh and Kenya and a tetanus booster campaign for adult women in Cambodia. In practice, the platform for integration could be adapted to local context and intervention need.

For each setting, we then identified the most efficient combination of pathogens as a case study, requiring that any intervention included targeted all of its pathogens (e.g., all *Plasmodium* species for malaria control), though not every intervention needed to be included. Then, we simulated all potential combinations of integrated interventions and identified the most efficient strategy integrating a VPD with at least one additional intervention in each location, guided by a multipathogen index calculated across all pathogens included in the strategy (**Figure 1**, Step 4). We required that each included intervention target all of its pathogens (e.g., all *Plasmodium* species for malaria control), though not every intervention needed to be included in the case study We compared the strategy guided by the multipathogen index to a strategy guided by a single VPD in each of the three settings, whereby single-pathogen strategies prioritized spatial clusters based on the prevalence of individuals who would benefit from an intervention delivering only vaccines. The purpose of this practical comparator analysis was twofold: first, to evaluate whether strategies using information from several pathogens could more efficiently accomplish several public health goals simultaneously compared to strategies focused on vaccines alone, and second, to assess the extent to which incorporating additional pathogens diminished the efficiency of a vaccine-focused intervention. We also compared a wealth-based strategy, which prioritized spatial clusters based on a multidimensional poverty index, agnostic to intervention need (**Supplementary Methods 2**). We estimated the proportion of clusters required to target 80% of potential beneficiaries under each strategy (the WHO Reaching Every District target for immunization coverage), and calculated 95% confidence intervals using bootstrapping with 1000 replicates and resampling spatial clusters with replacement.

We further evaluated the operational feasibility of the most efficient integrated intervention by assessing spatial aggregation of the multipathogen index (**Figure 1**, Step 5). We evaluated whether intervening on a coarser geographic unit, corresponding to subdistricts in Bangladesh and Kenya and provinces in Cambodia, provided similar efficiency gains as targeting at the spatial-cluster level. We assumed that targeting interventions at these coarser geographic levels may be more closely aligned with the geographic unit of practical intervention delivery.

### Characterizing populations experiencing high multipathogen index

Lastly, we evaluated the association between the multipathogen index calculated across all pathogens in each country and measures of poverty conditions using metadata collected at the household level. We estimated spatial-cluster level poverty using the framework for multidimensional poverty, a global standard that accounts for the share of a population that is impoverished and the intensity of their poverty^38,39^. The multidimensional poverty metric considers ten indicators spanning measures of health such as food security and child mortality, education, and living standards such as access to cooking fuel, electricity, and drinking water (**Supplementary Methods 2**).

To further characterize shared environmental and household factors associated with residing in high multipathogen index settings, we assessed the association between the multipathogen index and a set of predictor variables, capturing food security, sanitation, drinking water access, hygiene, electricity, cooking fuel, flooring and housing material, household assets (**Supplementary Methods 2**), and climate variables that can influence infectious disease risk, including mean annual temperature and precipitation^40^. For each location, predictor variables were aggregated to the spatial cluster level by taking the within-cluster mean of household-level indicators and environmental conditions. All household and environmental variables were rescaled to range between 0-100% for comparability.

For each location, we ranked cluster-level predictors by the proportion of between-tertile variance explained using a one-way ANOVA and retained the five most positively associated with the multipathogen index. Individual predictors indicating access to improved water, sanitation, and hygiene were removed from the Bangladesh and Kenya analyses given that study participants were randomized to these treatment groups, and thus access to improved conditions was evenly balanced among individuals.

### Ethics

The WASH Benefits trial (ClinicalTrials.gov identifier: NCT01590095) in Bangladesh and Kenya received ethical approval from multiple institutional review boards, including the Ethical Review Committee of the International Centre for Diarrhoeal Disease Research, Bangladesh (PR11063); the Scientific Ethics Review Unit (SERU) at Kenya Medical Research Institute, the Committee for the Protection of Human Subjects at the University of California, Berkeley (2011-09-3652); the Institutional Review Board at Stanford University (25863); and the Institutional Review Board at the University of California, San Francisco (22-36722). Ethical approval was previously obtained in Cambodia^30^.

## Results

### Evaluating candidate multipathogen indices across settings

We first compared several candidate multipathogen indices, including species richness, Shannon diversity index, Gini-Simpson index, and Rao’s quadratic index, for their ability to quantify overlapping public health intervention need (**Figure 2A**). We evaluated each index based on how well it identified efficient integrated strategies, whereby a more efficient strategy could reach more intervention beneficiaries by visiting fewer spatial locations. To quantify efficiency, we calculated the minimum number of spatial clusters needed to target at least 80% of all individuals in the population who would benefit from an intervention, corresponding to specified WHO targeting benchmarks^34,35^. We compared efficiency across the range of all possible intervention combinations including pathogens with potential for integration, defined here as pathogens that could be treated with current, large-scale public health interventions including vaccination, mass drug administration, or a malaria package, and for which ≥5% of the population could benefit from intervention, leading to 7 pathogens in the Bangladesh subregion (measles, rubella, diphtheria, tetanus, *Ascaris lumbricoides*, hookworm, and *Trichuris trichiura*), 8 in the Kenya subregion (measles, rubella, *Ascaris lumbricoides*, *Plasmodium falciparum, Plasmodium malariae, Plasmodium ovale,* Schistosomiasis, and *Taenia solium)* and 5 (tetanus, lymphatic filariasis, *Plasmodium falciparum, Plasmodium vivax,* and *Strongyloides stercoralis)* in Cambodia. This resulted in 99 combinations evaluated in the Bangladesh subregion, 219 in the Kenya subregion, and 16 in Cambodia (**Figure 2B**). We found that integrated strategies guided by Rao’s quadratic index consistently identified more efficient strategies than other multipathogen indices and additionally tended to outperform random prioritization (80% of all intervention beneficiaries targeted by visiting 80% of locations), with the exception of species richness in Kenya’s subregion.

**Figure 2.**
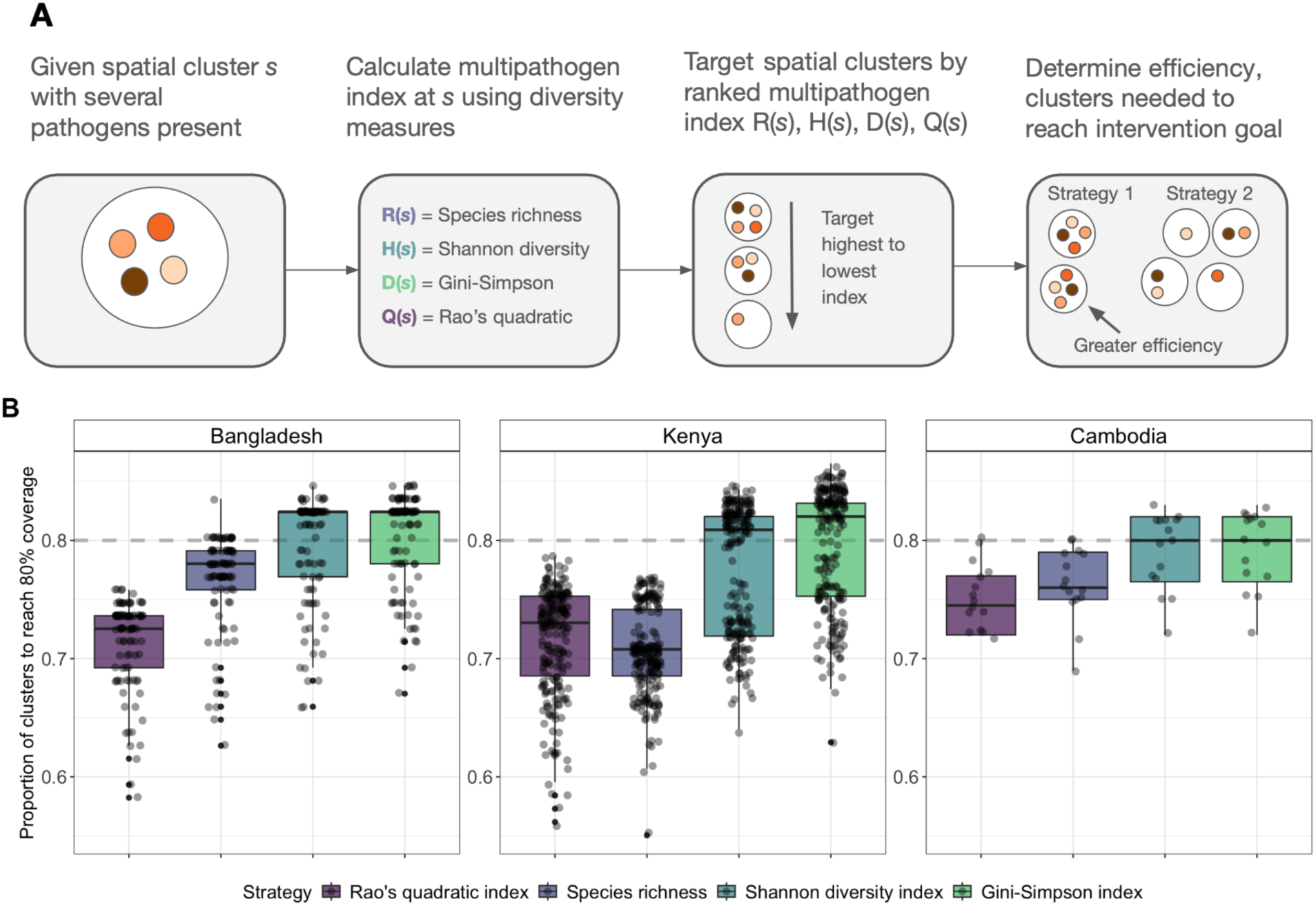
Comparing multipathogen indices and their ability to identify efficient strategies. **A)** Schematic demonstrating how multipathogen indices were applied to multipathogen surveillance data to guide spatial prioritization of integrated interventions. **B)** The efficiency of integrated strategies guided by several multipathogen indices, including species richness, Shannon diversity index, Gini-Simpson index, and Rao’s quadratic index. Efficiency is measured as the minimum number of spatial clusters that would need to be included in an integrated intervention to target a minimum of 80% of individuals who would benefit from an intervention for all pathogens considered, where more efficient strategies require fewer spatial locations. The boxplots correspond to the distribution of efficiency measured across all interventions, with individual points representing each unique intervention combination evaluated. The horizontal dashed gray line corresponds to the expected efficiency under random prioritization.

### Framework for identifying opportunities for integrated strategies

After finding that multipathogen strategies guided by Rao’s quadratic index tended to identify more efficient opportunities for integration, we then applied our proposed framework in each of the three settings where multipathogen surveillance data were available (**Figure 1**, Step 1). Having already identified pathogens with well-defined interventions and potential for integration (**Figure 1**, Step 2), we next defined existing vaccination campaigns as a realistic platform for integration (**Figure 1**, Step 3), corresponding to childhood measles immunization in the Bangladesh and Kenya subregions and a tetanus booster campaign for adult women in Cambodia. We simulated all possible combinations of interventions that included a vaccination campaign (**Figure 1**, Step 4). For each setting, we then identified the most efficient combination of pathogens as a case study, requiring that each intervention included target all of its pathogens (e.g., all *Plasmodium* species for malaria control), though not every intervention needed to be included. This resulted in the following integrated interventions: a strategy integrating childhood immunization campaigns (measles, rubella) with mass drug administration for soil-transmitted helminths (*Ascaris lumbricoides,* hookworm, and *Trichuris trichiura*) in the Bangladesh subregion, a strategy integrating a childhood immunization campaign (measles), malaria control (*Plasmodium falciparum, Plasmodium malariae, and Plasmodium ovale*), and mass drug administration (schistosomiasis) in the Kenya subregion, and a strategy integrating an adult booster vaccination campaign (tetanus), malaria control (*Plasmodium falciparum* and *Plasmodium vivax)*, and mass drug administration (lymphatic filariasis and *Strongyloides stercoralis*) in Cambodia (**Figure 3**).

**Figure 3.**
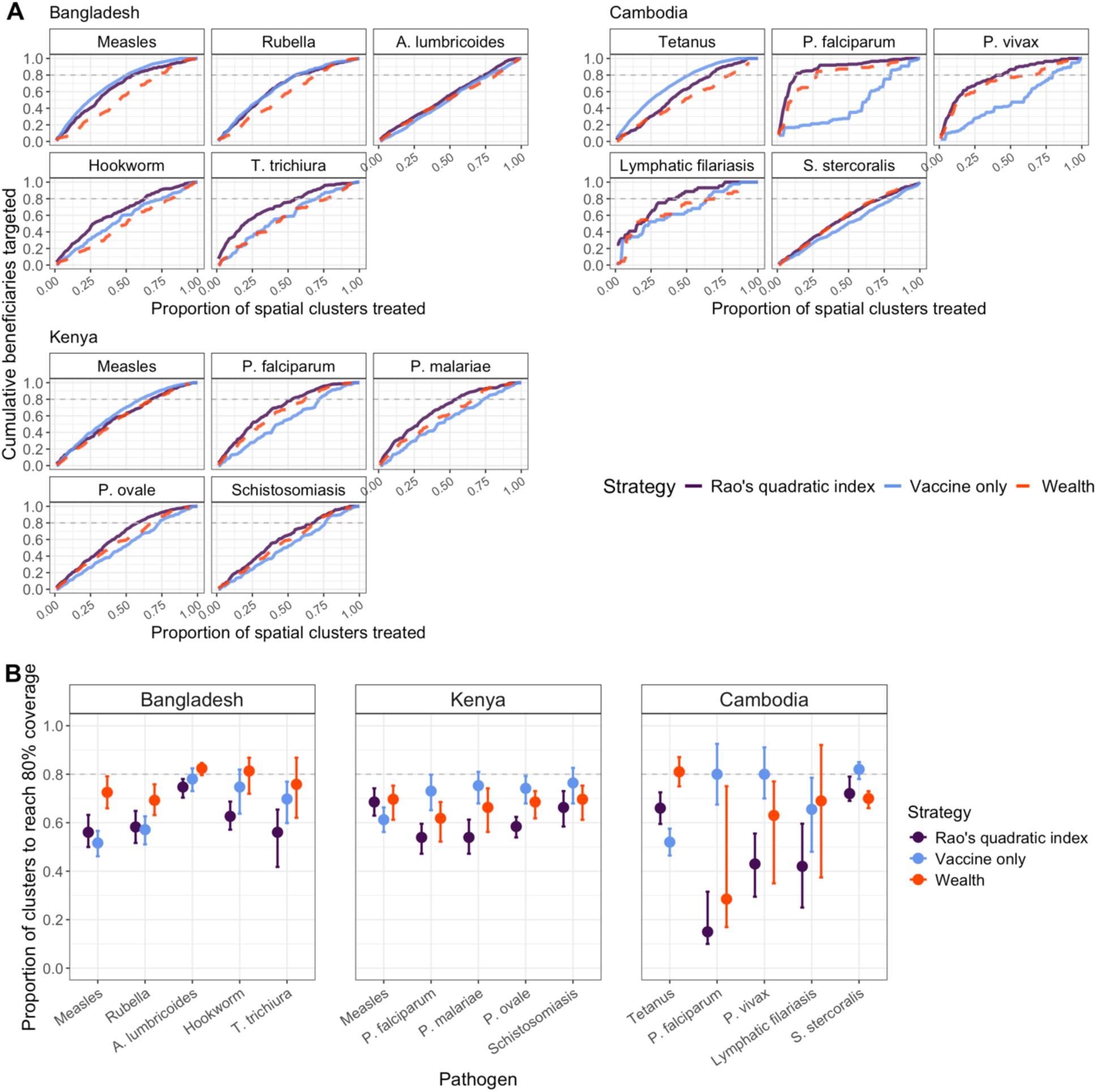
Efficient integrated strategies guided by multipathogen index against practical comparators. **A)** Efficiency curves shown for the most efficient integrated strategy guided by Rao’s quadratic index in each country. Plots show the cumulative fraction of beneficiaries (of vaccines, mass drug administration, or malaria control) as additional spatial clusters are reached for strategies guided by a multipathogen index (purple) against two practical comparator strategies guided by wealth (orange) and vaccination need (blue). **B)** The 95% confidence intervals for the proportion of clusters required to target 80% of beneficiaries for vaccine-guided, multipathogen-guided, and wealth-guided approaches, calculated using bootstrapping with 1000 replicates and sampling spatial clusters with replacement. The 95% confidence interval for the proportion of spatial clusters required to reach 80% coverage is consistently ≤80% (dashed gray line, expectation under random prioritization) across all settings for only the multipathogen-guided strategy, guided by Rao’s quadratic index.

Across each setting, we compared the efficiency of the integrated strategy guided by the multipathogen index (Rao’s quadratic index) to two practical comparator strategies guided by a single VPD and wealth. Across all settings, we found that a multipathogen index-guided strategy was more efficient on average than all practical comparator strategies targeting based on vaccine need or wealth (**Figure 3**). Compared to strategies targeting only vaccination need, incorporating information from other pathogens present in the system led to modest reductions in efficiency for VPD, while affording overall efficiency gains for additional pathogens (**Figure 3A**). We found that a multipathogen index-guided strategy reduced the number of spatial clusters needed to target 80% of all pathogens present by 4% (95% CI: 0-12%) in the Bangladesh subregion and 10% (95% CI: 3-17%) in the Kenya subregion compared to a strategy only using information on measles undervaccination to guide prioritization (**Figure 3B**). In Cambodia, a multipathogen index-guided strategy improved efficiency by 11% (95% CI: 5-22%) compared to a strategy guided only by tetanus undervaccination. The largest efficiency gains were observed for individual pathogens in Cambodia, with a multipathogen index-guided strategy reducing the number of spatial clusters needed by 83% (*Plasmodium falciparum*), 44% (*P. vivax*), 35% (lymphatic filariasis), and 11% (*Strongyloides stercoralis*), compared to 12-27% across individual pathogens in Kenya and 4-22% in Bangladesh. Relative to a wealth-based strategy, a strategy guided by the multipathogen index improved efficiency by 9% (95% CI: 3-19%) in the Bangladesh subregion, 3% (95% CI: -7-12%) in the Kenya subregion, and 11% (95% CI: 1-20%) in Cambodia. Strategies guided by the multipathogen index consistently outperformed random prioritization (equivalent to 80% of clusters required to reach 80% of beneficiaries) (**Figure 3B**).

Next, we studied spatial aggregation in the multipathogen index since geographic overlap between pathogens is a central concern for integrated intervention feasibility (**Figure 1**, Step 5). First, we mapped Rao’s quadratic index based on pathogens with tangible public health interventions in each location to visualize where overlapping intervention needs emerged (**Figure 4A**). The multipathogen index was spatially concentrated in the Northeastern regions of the Bangladesh subregion and Cambodia, and among the Southwestern region of Kenya, with moderate to high spatial autocorrelation detected in Bangladesh’s subregion (Moran’s I = 0.58), Kenya’s subregion (I = 0.56), and Cambodia (I = 0.26). Maps displaying Rao’s quadratic index at discrete sampling units are provided in **Supplementary Figure S3.** Second, we assessed the degree of spatial aggregation for the specific pathogens identified in the integrated strategy in **Figure 3**. We compared the efficiency of targeting at the spatial-cluster level to targeting coarser spatial units that may be more operationally feasible, corresponding to subdistricts in the Bangladesh (n = 18) and Kenya (n = 38) study regions and provinces in Cambodia (n = 22) (**Figure 4B**, spatial locations provided in **Supplementary Figure S4**). Across settings, we found that the efficiency of a multipathogen-guided strategy was almost unchanged when targeting interventions at the larger administrative unit, with slight efficiency losses for tetanus, *P. vivax*, and lymphatic filariasis in Cambodia (**Figure 4B**).

**Figure 4.**
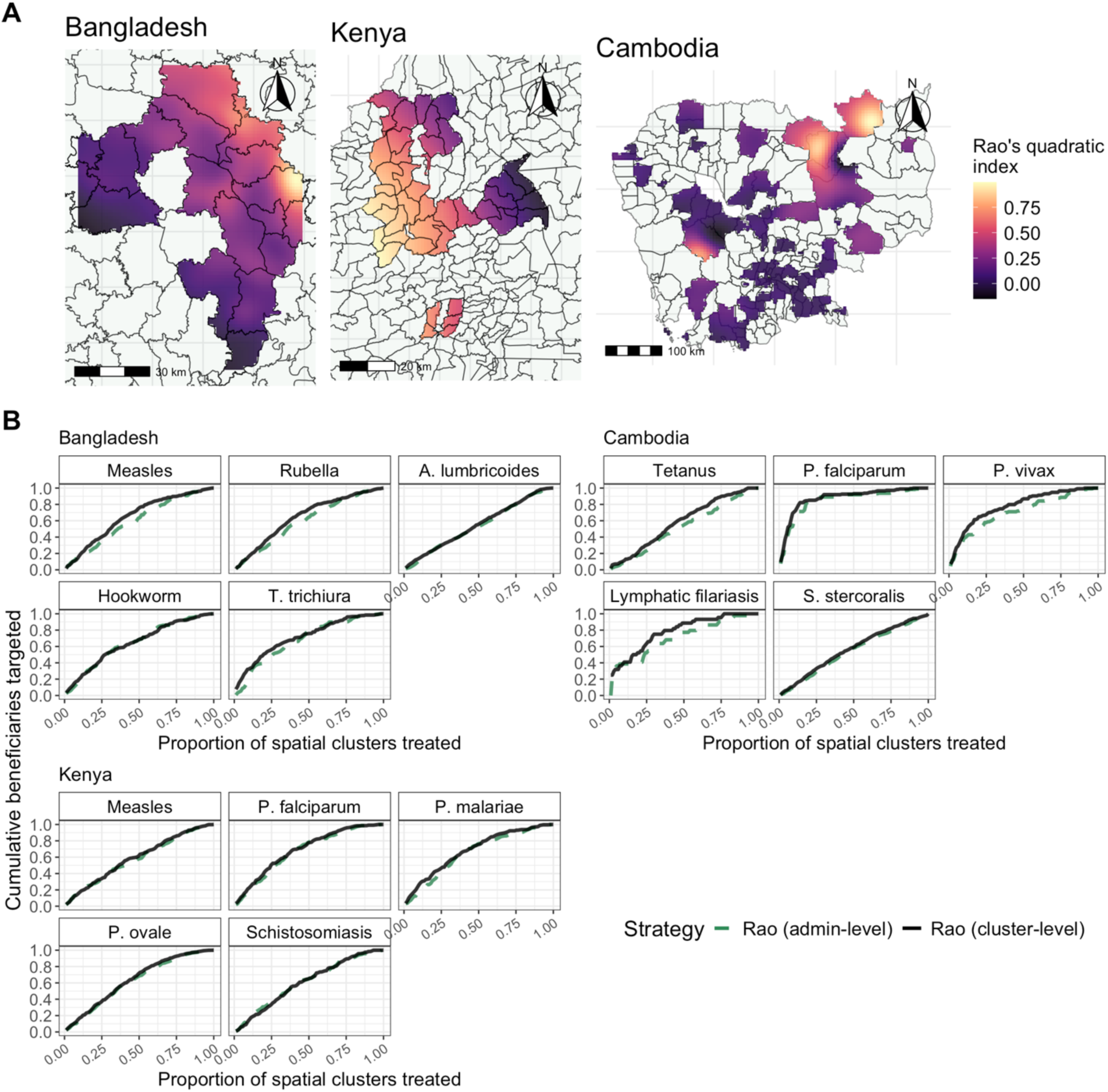
Spatial aggregation of multipathogen index across settings. **A)** Smoothed multipathogen index (Rao’s quadratic index) calculated at the spatial-cluster level and mapped in each setting (n = 90 spatial clusters in Bangladesh, n = 89 in Kenya, and n = 100 in Cambodia, respectively). A geostatistical model with semi-parametric smoothing via a Matérn covariance structure was fit to spatial-cluster-level multipathogen index values, assuming a Gaussian process, as a function of latitude and longitude. Smoothed predictions were masked to only display administrative units where there was at least one spatial cluster present. Lines overlaying the map indicate subdistrict boundaries. Multipathogen index values for discrete sampling units are provided in **Supplementary Figure S3. B)** Using the most efficient integrated intervention in each setting as described above, we compared the efficiency of a strategy that prioritized locations based on multipathogen index calculated at the spatial cluster level (black, solid line) and a coarser spatial unit (green, dashed line). The coarser spatial unit corresponded to subdistricts in Bangladesh’s subregion (n=18) and Kenya’s subregion (n = 38), and provinces in Cambodia (n =22).

### Characterizing populations experiencing high multipathogen index

Finally, we evaluated whether the multipathogen index, calculated over pathogens with tangible interventions in each setting, may reflect measures of population well-being. We found that a cluster-level multipathogen index was significantly correlated with independent measures of multidimensional poverty in Bangladesh’s subregion (Spearman’s correlation coefficient ρ = 0.25, p-value = 0.025), Kenya’s subregion (ρ = 0.51, p-value < 0.001) and Cambodia (ρ = 0.59, p-value < 0.001) (**Figure 5A**). In each location, we then identified the five household and environmental factors that explained the highest proportion of between-tertile variance in the multipathogen index, considering household conditions, access to improved Water-Sanitation-Hygiene infrastructure, environmental risk factors, and other measures of well-being (**Figure 5B).** Across settings, factors related to housing quality, including flooring, housing material, and household assets, were consistently associated with a high multipathogen index. Additional environmental risk factors were associated with the multipathogen index, including high precipitation in Bangladesh and hotter temperatures in Kenya subregions. In Cambodia, clusters with the highest multipathogen index had greater hygiene and sanitation deprivation, accounting for 16 and 17% of the variation between tertiles of the multipathogen index, respectively.

**Figure 5.**
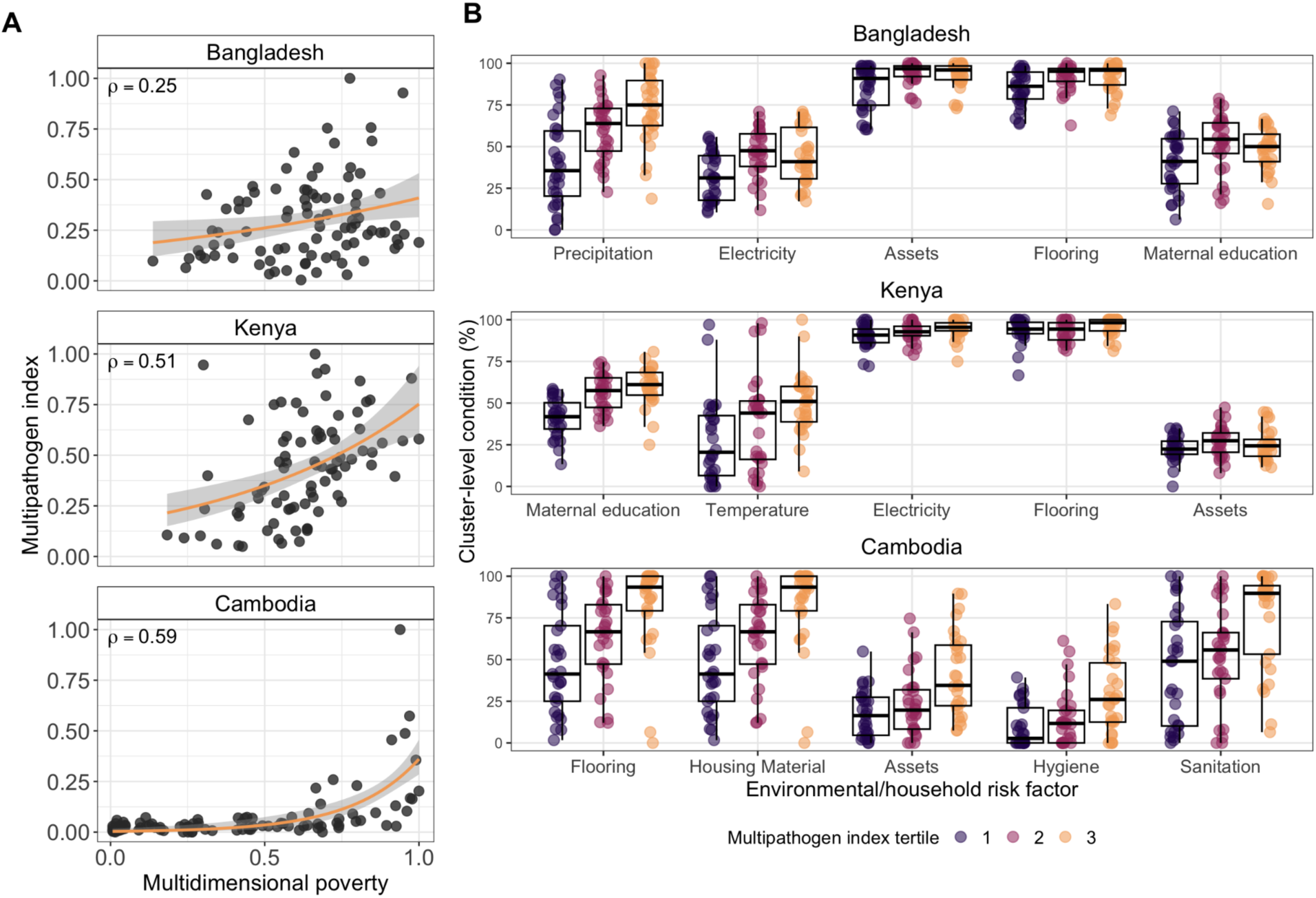
Comparing multipathogen indices to wealth and environmental/household conditions. **A)** Multidimensional poverty plotted against Rao’s quadratic index, calculated over the full set of pathogens, with each point corresponding to a single spatial cluster, by location. **B)** For each location, we ranked cluster-level risk factors by the proportion of between-tertile variance explained using a univariate ANOVA and retained the five most positively associated with the multipathogen index. Boxplots display the distribution of each identified cluster-level condition within each tertile of the multipathogen index. Household indicators measure the percentage of each community that is deprived of the indicator, whereby higher deprivation generally corresponds to higher multipathogen index tertiles, consistent with greater pathogen co-occurrence in more vulnerable spatial clusters. Climate variables (temperature and precipitation) were measured continuously and standardized on the 0-100% scale.

## Discussion

We developed a framework to use multipathogen surveillance data to identify opportunities for more efficient integrated intervention strategies. Across three diverse settings with varied intervention needs, surveyed populations, and diagnostic assay platforms, we demonstrated that applying a multipathogen index (Rao’s quadratic index) consistently identified more efficient integrated strategies compared with other common theoretical measures of diversity, and was able to achieve multipathogen targeting simultaneously across a range of intervention goals (**Supplementary Figure S2**). The performance of Rao’s quadratic index over other measures of diversity in this analysis follows from the way it integrates information across pathogens: while the Shannon diversity index is more sensitive to rare quantities and the Gini-Simpson index places greater weight on common quantities and both are compositional quantities^41^, Rao’s quadratic index prioritizes spatial locations with several pathogens present and high pairwise prevalences, enabling for more efficiency gains in this application. Using Rao’s quadratic index as a multipathogen index, the proposed framework identified efficient and feasible multipathogen-guided strategies for integrated interventions. We found that these strategies were more efficient than practical comparator strategies guided by a single VPD or wealth-based targeting, affording efficiency gains between 3-11% across settings. Given that health intervention financing can often be largely attributed to administrative, travel, and personnel costs^42,43^, even modest reductions in the number of spatial locations required to visit in an intervention could lead to large cost savings^44^. Finally, we found that overlapping intervention needs were geographically concentrated in all three case studies, suggesting that integrated strategies guided by a multipathogen index will likely be logistically feasible and align with administrative units at the relevant scale of decision-making.

We found that the efficiency of specific integrated interventions depends on the individual diseases included and their underlying spatial distributions. In general, the index tended to identify efficient strategies when pathogens had intermediate prevalences and were focally concentrated in space. Although the three case studies presented here were based on immunization platforms, removing this constraint revealed more efficient integrated strategies for other diseases. For example, an integrated strategy in Cambodia not tied to vaccination was able to target 80% of all focally concentrated malaria and lymphatic filariasis cases by reaching 39% of spatial locations, compared to 72% for the strategy presented in the main text. Conversely, for pathogens that infect high proportions of the population or are evenly distributed geographically, an integrated strategy may offer little advantage over pathogen-focused interventions. Efficiency gains over vaccine-motivated strategies were modest in the Bangladesh subregion, largely because *Ascaris lumbricoides* was evenly distributed across study districts and demonstrated high population-level prevalence (36.6%), thus requiring a high number of spatial locations to reach 80% of intervention need regardless of prioritization strategy.

By considering a range of hypothetical interventions, the proposed framework allows the user to determine when a multipathogen-motivated strategy offers large efficiency gains or when a traditional single pathogen-motivated strategy may be sufficient, enabling more informed programmatic decision-making. For instance, vaccine-based targeting in Bangladesh and wealth-based targeting in Kenya achieved similar efficiency as multipathogen prioritization, illustrating how multipathogen data can identify settings where simpler strategies suffice, and demonstrating that the most parsimonious strategy may differ between settings. We also note that while the framework outlines a stepwise workflow, it could be adapted to incorporate iterative or nonlinear feedback; for instance, evaluating spatial aggregation and feasibility of interventions in Step 5 could inform or refine which pathogens are considered as having potential for integration in Step 2.

This framework aligns with the World Health Organization’s 2021-2030 roadmap to prevent, control, and eliminate 20 diseases and broader disease groups, which includes the integration of disease interventions as a core strategy^8^. Reductions in global health funding have exacerbated the need for more efficient, cross-cutting interventions, with funding cuts particularly disrupting NTD control and elimination efforts^10^. When multipathogen surveillance data is available at scale, our findings highlight the potential to integrate NTD and other interventions into programs with more stable funding, such as vaccine programmes, at a minimal cost to the efficiency of the core vaccination campaigns. Integrated delivery is already being implemented in areas with overlapping intervention needs. For instance, an integrated measles campaign in Ethiopia reached 18.5 million children in 2025, jointly providing deworming medication, Vitamin A supplementation, and other healthcare services alongside routine measles immunization, and the Tanzanian Ministry of Health has coordinated the integrated delivery of immunization and neglected tropical diseases programs, reaching over 20 million people in recent campaigns^45,46^. Safety trials are also being conducted across settings to ensure integrated treatment regimens are well tolerated. For example, Zanzibar recently integrated the delivery of praziquantel, ivermectin, and albendazole within coordinated deworming campaigns to concurrently target schistosomiasis, lymphatic filariasis, onchocerciasis, and soil-transmitted helminths^47^, and in Ethiopia, trials have evaluated simultaneous delivery of albendazole, ivermectin, and azithromycin to target lymphatic filariasis, onchocerciasis, trachoma, and soil-transmitted helminths^48^. These integration efforts are increasingly viable due to established cross-pathogen support structures such as the WHO Regional Office for Africa’s Expanded Special Project for Elimination of Neglected Tropical Diseases (ESPEN), which provides national programs with technical, resource-mobilization, and supply-chain support to coordinate treatment across co-endemic diseases^49^. Our results suggest that multipathogen data could strengthen these efforts by providing opportunities to systematically identify where intervention needs co-occur and which interventions can be most efficiently combined.

Importantly, these multipathogen data are increasingly available at scale, due to growing investment from collaborations spanning national ministries of health, universities, nonprofits, and industry. A joint initiative between the Pan American Health Organization, the World Health Organization, and the US CDC recently developed a framework for integrated serological surveillance in the Americas^23^, which has supported the expansion of large-scale multiplex serosurveys in Brazil, Mexico, Paraguay, Guyana, and Guatemala^28,29,50^. Several individual countries, including Nigeria, Kenya, Zambia, Samoa, and Tonga, have recently implemented or received grants to conduct integrated national serosurveillance surveys, and several are investing in sustainable in-country laboratory and diagnostic capacity^24–27,51–54^. For example, the Kenya Multi-Site Serosurveillance (KEMIS) collaboration has brought together stakeholders from the National AIDS and STIs Control Program and the National Malaria Control Program to support the development of nationally representative serosurveillance efforts^55^. Our analysis supports the continued investment in priority multipathogen surveillance to identify new opportunities for more efficient integration under resource constraints. While the framework is agnostic to the format of surveillance data used, requiring only spatially disaggregated prevalence estimates, platforms such as multiplex serosurveillance, which can measure tens of antigens simultaneously^3,7^, may improve efficiency and cost-effectiveness.

In each setting examined, we found that independent measures of poverty were strongly correlated with a multipathogen index. Despite this, multipathogen-motivated strategies achieved greater efficiency than wealth-based strategies on average in all settings. This suggests that a serological assay may be a useful tool for distinguishing locations where intervention needs and poverty diverge, which could arise due to a variety of environmental or infrastructural factors. For example, animal ownership was associated with improved access to household assets and wealth in the Kenya subregion, yet agricultural occupations involving animal ownership can also predispose individuals to increased infectious disease risk due to frequent contact with animals and other environmental exposures, which could potentially explain how high multipathogen indices could be observed among populations with higher socioeconomic status^56^.

This contrasts with Cambodia, where high multipathogen indices were almost exclusively concentrated in locations with the highest impoverishment, focally concentrated in the Northeastern region where densely wooded areas facilitate mosquito-breeding grounds and remoteness presents barriers to improved socioeconomic conditions^57^. Given the nuanced relationship between impoverishment and high multipathogen indices exhibited across settings, our results suggest that a multipathogen index could complement existing, well-validated poverty measures as a biomarker-based approach to identifying impoverished populations with the highest public health need^58–61^. Because multipathogen surveillance data can provide a biologically grounded window into a community’s unique landscape of disease risk, it can shed light on where communities face multiple infectious disease threats or experience gaps in healthcare service delivery. This information could be particularly useful for informing where to prioritize healthcare strengthening efforts that can improve resilience across diverse disease systems.

The approach has several limitations. First, the framework simplifies the integration process and does not consider important aspects of intervention design such as the underlying seasonality of disease, the cost or feasibility of intervention, the mode of delivery, or the target population. Some single-pathogen interventions target narrow age groups (e.g., childhood immunization campaigns) or are delivered only during specific periods, so the surveyed population must align with the intervention’s target population in both age and timing. Since the framework evaluates all potential combinations of integrated strategies, in principle, it can incorporate logistical constraints to restrict the analysis to interventions with viable, context-specific integration potential. Second, our analysis derives prevalence estimates for many pathogens by using multiplex IgG data and established seropositivity cutoffs. Serology measures different timescales of infection depending on pathogen-specific immunological dynamics, with some antibody responses persisting months or even years following active infection for some antigens^62^. Given that the majority of our study populations comprised young children <30 months in the Bangladesh and Kenya subregions, seropositivity reflects recent pathogen exposure, but if designing interventions to target older populations, it may be useful to augment serological surveillance data with other measures of infection or ensure that antigens used represent recent exposure and timely intervention need. Finally, while the serosurvey was nationally representative in Cambodia, enrollment in the Kenya and Bangladesh subregions was based on pregnancy timing and children were measured over an extended period of time that may coincide with underlying seasonality patterns, and thus may not reflect the broader community intervention needs; improvements in national serosurveillance capacity will enable more representative and systematic random sampling of populations to inform integration in the future.

This analysis largely relied on surveillance data from large-scale serological studies paired with stool-based microscopy for soil-transmitted helminths in the Bangladesh and Kenya subregions, providing a realistic example of how different diagnostics could be combined to inform integration. While our framework can accommodate prevalence data generated from across surveillance platforms, multipathogen surveillance platforms may be more logistically challenging to implement when there are pathogen-specific diagnostic preferences or distinct funding structures and divisions supporting single-pathogen management. For example, it is common for malaria interventions to be designed around incidence/prevalence data from rapid diagnostic tests rather than seroprevalence^63^. More broadly, our approach requires established and sustainable surveillance infrastructure. Residual sera are often routinely available through healthcare systems and could be leveraged to support multipathogen surveillance efforts^64^, and many large-scale, nationally representative serosurveillance efforts are underway, but operational feasibility and cost may remain key barriers to implementation in settings with overlapping intervention needs. Finally, we used an intervention goal of 80% of intervention beneficiaries targeted across pathogens in this analysis, aligning with the WHO Reaching Every District initiative^34,35^. National programmes may set different targeting goals dependent on the disease or abide by WHO pathogen-specific guidelines for treatment. In this case, the framework could be extended to measure efficiency towards meeting pathogen-specific targeting goals.

## Conclusion

These findings show that multipathogen surveillance can identify opportunities to deliver integrated interventions more efficiently, with Rao’s quadratic index providing a theoretical basis for identifying locations where intervention needs are spatially aggregated. This framework can be applied across diverse settings and diagnostic platforms and may jointly serve as a biomarker-based indicator of impoverishment for detecting where populations experience multilayered health disparities. As health systems continue to face constrained resources, integrating multipathogen surveillance into intervention planning could enable for more efficient coordinated approaches to disease control.

## Data Availability

All data and code used in the study is publicly available at: https://github.com/sbents/multipathogen

## Acknowledgements

The authors thank Gretchen Cooley and Diana L. Martin at the US Centers for Disease Control and Prevention for their multiplex serology antigen bead coupling, protocols, laboratory training and ongoing support.

## Code and Data Availability

All data used in the study is publicly available and code used for all analyses is publicly available at: https://github.com/sbents/multipathogen.

