## Supplementary files for "A framework for guiding integrated disease control measures through multipathogen surveillance"

**Supplementary Figures and Tables for:** A framework for guiding integrated disease control measures through multipathogen surveillance

**Table S1. Recommended treatment and control strategies by pathogen.** Summary of current guidance on the control of study pathogens. We reported treatment recommendations only for the pathogens for which at least ≥ 5% of the population could benefit from associated intervention across settings.

| **Pathogen** | **Intervention guidelines** |
|---|---|
| *Ascaris lumbricoides* | Administration with single dose albendazole or mebendazole in endemic areas, annual administration if prevalence ≥ 20% and biannual if prevalence ≥ 50%[1]. |
| *Trichuris trichiura* | Administration with single dose albendazole or mebendazole in endemic areas, annual administration if prevalence ≥ 20% and biannual if prevalence ≥ 50%[1]. |
| Hookworm | Administration with albendazole or mebendazole in endemic areas, annual administration if prevalence ≥ 20% and biannual if prevalence ≥ 50%[1]. |
| Measles | Prevention via measles-containing vaccine (e.g., two doses of MMR) in childhood[2]. |
| Rubella | Prevention via rubella-containing vaccine (e.g., two doses of MMR) in childhood[2]. |
| Tetanus | Prevention via tetanus-toxoid containing vaccine (e.g., pentavalent DTP-HepB-Hib vaccine) administered in childhood[2]. |
| Diphtheria | Prevention via diphtheria-containing vaccine (e.g., pentavalent DTP-HepB-Hib vaccine) administered in childhood[2]. |
| Schistosomiasis | Administration with single dose of praziquantel where prevalence ≥ 10%[1,3]. |
| *Taenia solium* | Administration with praziquantel, niclosamide, or albendazole in endemic areas[4], Neurocysticercosis treated with anthelminthics in combination with corticosteroids[5]. |
| *Plasmodium falciparum* | Mass drug administration with artemisinin-based combination therapy (ACT) and malaria vaccination with RTS,S/AS01 or R21/Matrix-M in endemic areas[6]. Household-level vector control efforts including insecticide-treated nets and indoor residual spraying[6]. |
| *Plasmodium malariae* | Mass drug administration with ACT and household-level vector control efforts including insecticide-treated nets and indoor residual spraying[6]. |
| *Plasmodium ovale* | Mass drug administration with ACT and household-level vector control efforts including insecticide-treated nets and indoor residual spraying[6]. |

| | |
|---|---|
| *Plasmodium vivax* | Mass drug administration with artemisinin-based combination therapy (ACT) and household-level vector control efforts including insecticide-treated nets and indoor residual spraying[6]. |
| *Strongyloides stercoralis* | Administration with single dose of ivermectin if prevalence ≥ 10%[7]. |
| Lymphatic filariasis | Administration with albendazole, ivermectin plus albendazole, diethylcarbamazine (DEC) plus albendazole, or IDA (ivermectin, albendazole, and DEC)[8]. |

**Table S2. Pathogens evaluated by multipathogen surveillance in each setting**. The full number of pathogens evaluated by each surveillance platform, the subset of priority pathogens for which ≥5% of the population is impacted, and the list of priority pathogens for which a tangible intervention exists. All main text analyses concentrate on only pathogens in the third column.

| Location | All pathogens evaluated | Pathogens for which ≥5% of the population is impacted | Pathogens with a tangible intervention for which ≥5% of the population could benefit from its intervention, used in main analysis |
|---|---|---|---|
| Bangladesh subregion | Serological dataset: *Entamoeba histolytica, Campylobacter*, chikungunya, cholera, *cryptosporidium*, dengue, diphtheria, Giardia, LT-ETEC , measles, norovirus, onchocerciasis, *Plasmodium falciparum, Plasmodium malariae, Plasmodium ovale, Plasmodium vivax,* rubella, *Salmonella,* schistosomiasis, *Shigella, Strongyloides, Taenia solium*, tetanus, trachoma, Zika<br><br>Stool-based microscopy: *Ascaris lumbricoides, Trichuris trichiura,* and hookworm | *Cryptosporidium,* norovirus, *Campylobacter*, *Salmonella, Giardia*, rubella, *Shigella*, measles, tetanus, diphtheria, cholera, LT-ETEC, *Ascaris lumbricoides, Trichuris trichiura,* hookworm | VPD:<br>1. Measles<br>2. Rubella<br>3. Diphtheria<br>4. Tetanus<br><br>MDA (albendazole):<br>5. *Ascaris lumbricoides*<br>6. *Trichuris trichiura*<br>7. Hookworm |
| Kenya subregion | Serological dataset: *Entamoeba histolytica, Campylobacter*, chikungunya, cholera, *cryptosporidium*, dengue, diphtheria, Giardia, LT-ETEC , measles, norovirus, onchocerciasis, *Plasmodium falciparum, Plasmodium malariae, Plasmodium ovale, Plasmodium vivax,* rubella, *Salmonella,* schistosomiasis, *Shigella, Strongyloides, Taenia solium*, tetanus, trachoma, Zika<br><br>Stool-based microscopy: *Ascaris lumbricoides, Trichuris trichiura,* and hookworm | *Ascaris lumbricoides, Taenia solium, Entamoeba histolytica, cryptosporidium*, *Plasmodium falciparum*, Schistosomiasis, *Plasmodium ovale, Plasmodium malariae, Campylobacter, Salmonella*, Giardia, rubella, measles, *Shigella*, norovirus, cholera, LT-ETEC | VPD:<br>1. Rubella<br>2. Measles<br><br>MDA (e.g., albendazole or praziquantel):<br>3. *Ascaris lumbricoides*<br>4. *Taenia solium*<br>5. Schistosomiasis<br><br>Malaria:<br>6. *Plasmodium falciparum*<br>7. *Plasmodium malariae*<br>8. *Plasmodium ovale* |
| Cambodia | Serological dataset: lymphatic filariasis, *Plasmodium falciparum, Plasmodium vivax, Strongyloides stercoralis*, *Taenia solium, Toxoplasma gondii,* Tetanus | lymphatic filariasis, *Plasmodium falciparum, Plasmodium vivax, Strongyloides stercoralis*, *Toxoplasma gondii,* Tetanus | VPD:<br>1. Tetanus<br><br>MDA (ivermectin):<br>2. *Strongyloides stercoralis*<br>3. Lymphatic filariasis<br><br>Malaria:<br>4. *Plasmodium falciparum*<br>5. *Plasmodium vivax* |

**Figure S1. Pathogen-specific prevalences by setting.** Distribution of the cluster-level proportion of the population that would benefit from interventions for each of the study pathogens in the Bangladesh and Kenya subregions and Cambodia. *Ascaris lumbricoides, Trichuris trichiura,* and hookworm prevalence were measured using stool-based microscopy and remaining pathogens were measured via IgG in a multiplex serological bead assay.

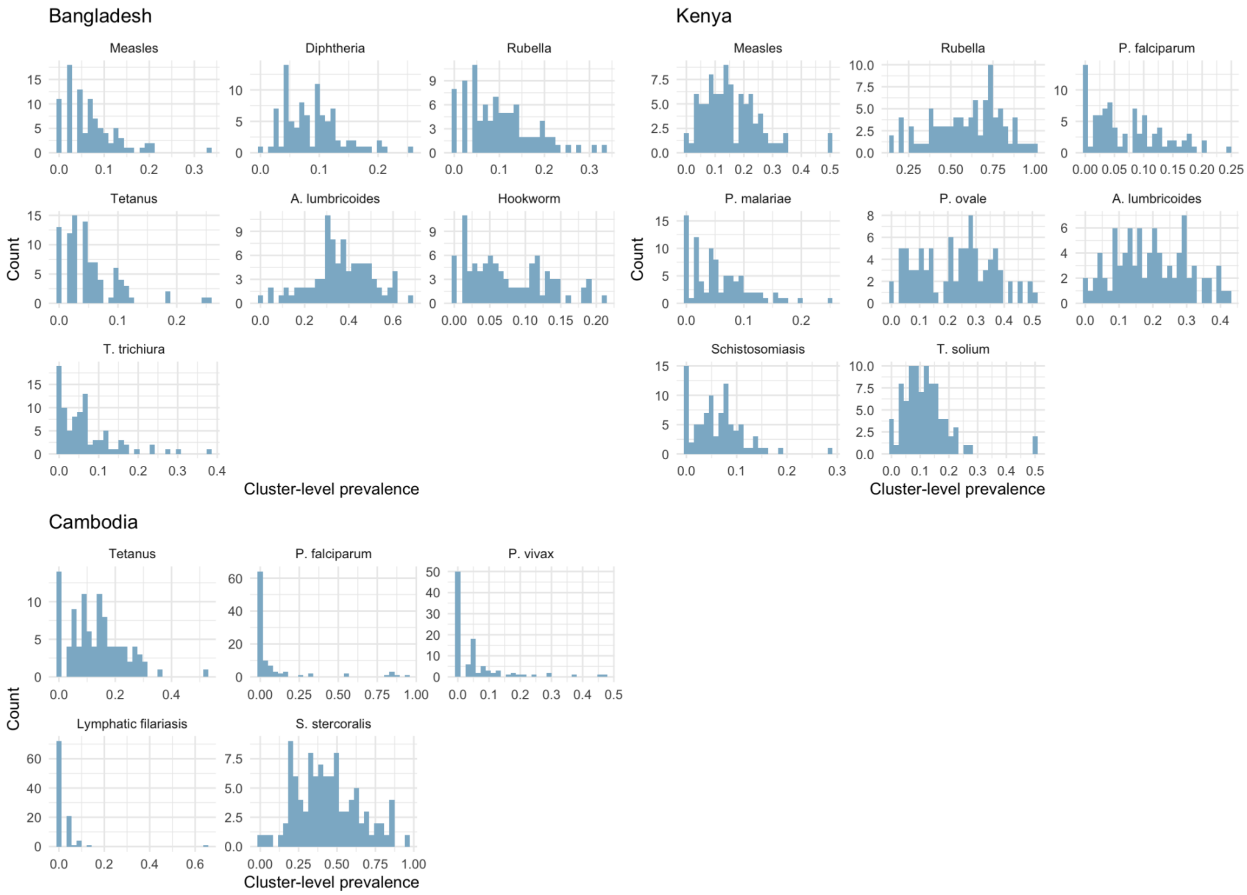

**Figure S2. Efficiency of integrated strategies across targeting goals.** We evaluated the efficiency of integrated interventions across different target thresholds, where efficiency was defined as the proportion of spatial clusters needed to reach the targeting goal of 50% (top) or 90% (bottom) of intervention beneficiaries across all pathogens included in each possible integrated strategy combining interventions for at least three pathogens. The multipathogen strategy guided by Rao's quadratic index consistently outperformed other diversity metrics across settings and targeting thresholds.

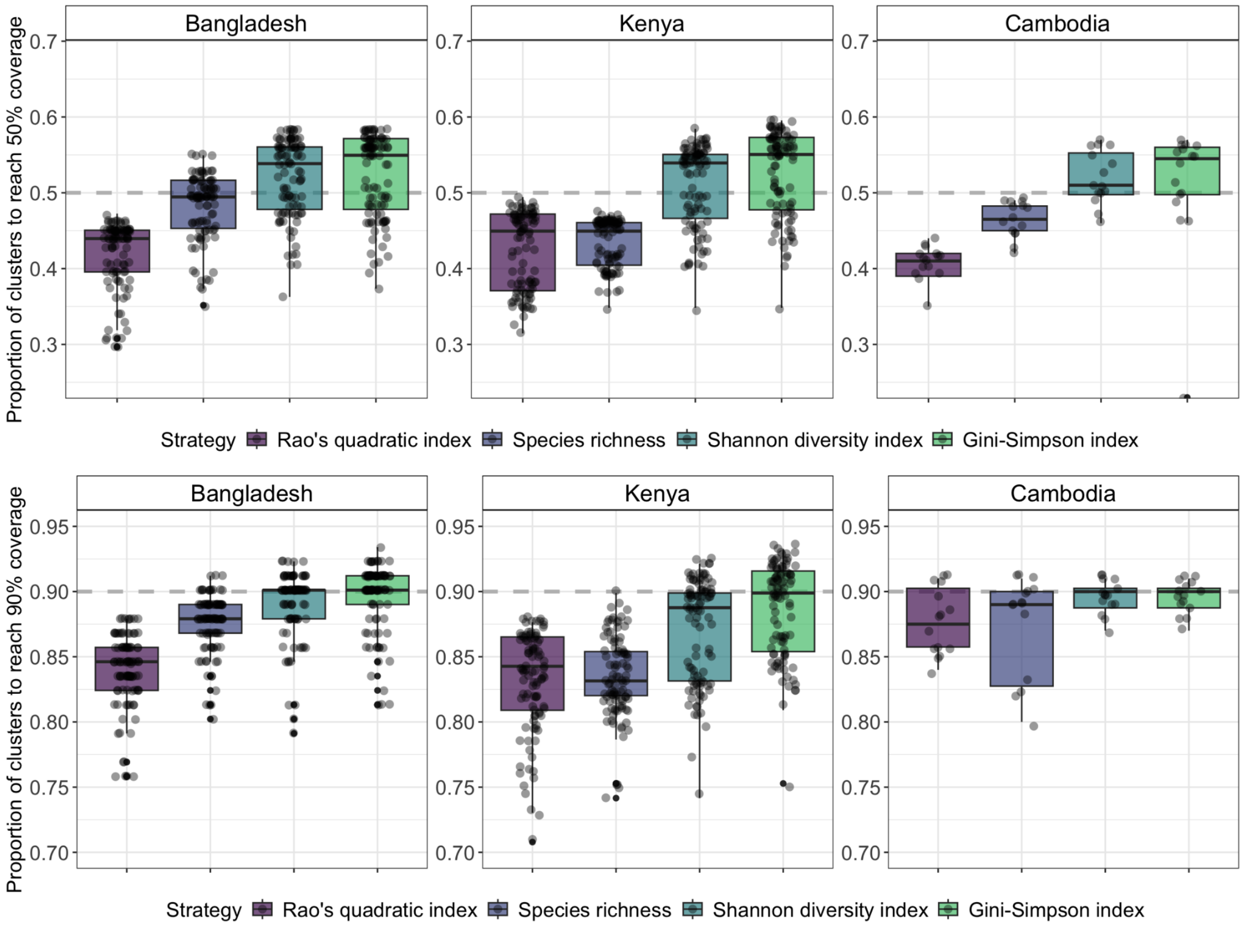

**Figure S3. Multipathogen index maps across settings.** Individual points correspond to spatial clusters and are colored by the value of Rao's quadratic index, calculated over all pathogens (n = 90 spatial clusters in Bangladesh, 89 in Kenya, and 100 in Cambodia), where lighter colors correspond to higher values of the multipathogen index.

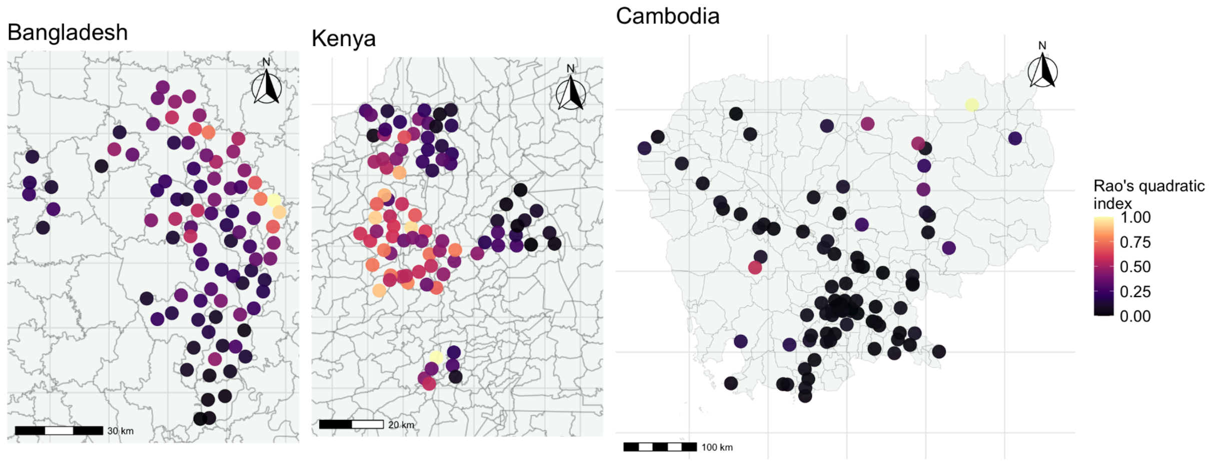

**Figure S4. Multipathogen index calculated at a broader administrative unit.** Multipathogen index (Rao's quadratic index) calculated at the subdistrict level in Bangladesh (n =18) and Kenya (n =38) and at the province level in Cambodia (n =22). Triangles show administrative units, where lighter colors correspond to higher values of the multipathogen index. Lines indicate subdistrict boundaries.

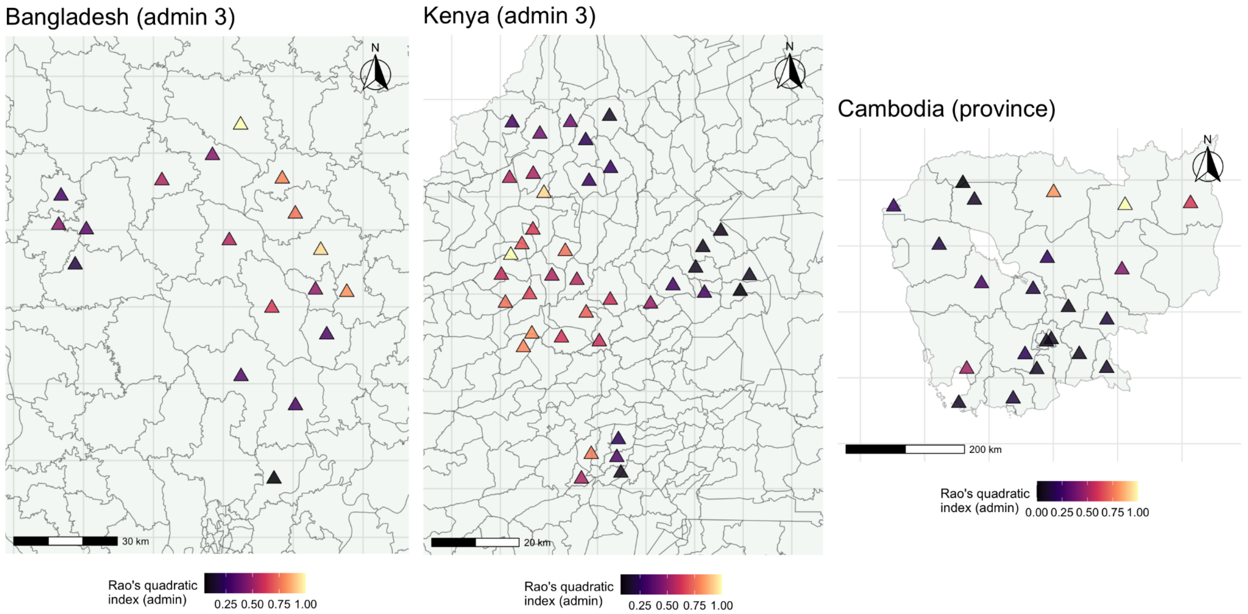

### Supplementary Methods

**Supplementary Methods 1.** In the main analysis, we used Rao's quadratic index to evaluate every possible combination of three or more pathogens in each setting. For each combination, we set $d_{i,j} = 1$ for all pairs of pathogens included in the integrated intervention and $d_{i,j} = 0$ otherwise, so that every included pair contributed equally to Q(s), regardless of whether the pathogens could be targeted by the same intervention or required separate, appended strategies. Because this approach treats all pathogens within an integrated intervention as jointly targetable, it does not capture the additional operational efficiency of interventions that can target several pathogens simultaneously.

Here, we instead defined $d_{i,j} = 1$ only when two pathogens share an exact intervention, and $d_{i,j} = 0$ otherwise (based on recommended interventions in **Supplementary Table S1**). The choice of $d_{ij}$ depends on the intended use. Setting $d_{i,j} = 1$ for all pairs is most appropriate when the goal is to identify spatial overlap in burden where relevant interventions are available, irrespective of delivery platform, consistent with the main analysis. Setting $d_{i,j} = 1$ only for pathogens with a shared intervention may be more appropriate when the goal is also to prioritize areas where a single intervention can address several pathogens simultaneously, emphasizing operational feasibility alongside spatial co-occurrence.

We evaluated the same integrated combinations of interventions using both interpretations of $d_{i,j}$ to assess whether spatial cluster prioritization shifted under different specifications. The shared-intervention matrices for each setting were defined as:

Kenya subregion

| | Rubella | Measles | *Ascaris lumbricoides* | *Taenia solium* | Schistosomiasis | *P. falciparum* | *P. malariae* | *P. ovale* |
|---|---|---|---|---|---|---|---|---|
| Rubella | -- | 1 | 0 | 0 | 0 | 0 | 0 | 0 |
| Measles | 1 | -- | 0 | 0 | 0 | 0 | 0 | 0 |
| *Ascaris lumbricoides* | 0 | 0 | -- | 1 | 0 | 0 | 0 | 0 |
| *Taenia solium* | 0 | 0 | 1 | -- | 1 | 0 | 0 | 0 |
| Schistosomiasis | 0 | 0 | 0 | 1 | -- | 0 | 0 | 0 |
| *P. falciparum* | 0 | 0 | 0 | 0 | 0 | -- | 1 | 1 |
| *P. malariae* | 0 | 0 | 0 | 0 | 0 | 1 | -- | 1 |
| *P. ovale* | 0 | 0 | 0 | 0 | 0 | 1 | 1 | -- |

Bangladesh subregion

| | Rubella | Measles | Diphtheria | Tetanus | *Ascaris lumbricoides* | *Trichuris trichiura* | Hookworm |
|---|---|---|---|---|---|---|---|
| Rubella | -- | 1 | 0 | 0 | 0 | 0 | 0 |
| Measles | 1 | -- | 0 | 0 | 0 | 0 | 0 |
| Diphtheria | 0 | 0 | -- | 1 | 0 | 0 | 0 |

| | | | | | | | |
|---|---|---|---|---|---|---|---|
| Tetanus | 0 | 0 | 1 | -- | 0 | 0 | 0 |
| *Ascaris lumbricoides* | 0 | 0 | 0 | 0 | -- | 1 | 1 |
| *Trichuris trichiura* | 0 | 0 | 0 | 0 | 1 | -- | 1 |
| Hookworm | 0 | 0 | 0 | 0 | 1 | 1 | -- |

Cambodia

| | Tetanus | *Strongyloides stercoralis* | Lymphatic filariasis | *P. falciparum* | *P. vivax* |
|---|---|---|---|---|---|
| Tetanus | -- | 0 | 0 | 0 | 0 |
| *Strongyloides stercoralis* | 0 | -- | 1 | 0 | 0 |
| Lymphatic filariasis | 0 | 1 | -- | 1 | 1 |
| *P. falciparum* | 0 | 0 | 1 | -- | 1 |
| *P. vivax* | 0 | 0 | 1 | 1 | -- |

For each integrated combination, we ranked spatial clusters by Q(s) computed under each matrix and calculated the proportion of spatial clusters that must be targeted to reach 80% of all intervention beneficiaries in that specific combination, consistent with the main analysis. Integrated interventions containing no pair of pathogens with a shared intervention were excluded from this comparison, because Q(s) = 0 in every cluster under the shared-intervention matrix and therefore the cluster ranking would be undefined.

Prioritizing clusters using shared-intervention matrices generally required a larger proportion of spatial clusters to reach 80% of burden, with a median increase of 5.5% in Bangladesh, 6.5% in Cambodia, and 2.2% in Kenya (**Supplementary Figure S5**). The shared-intervention approach required more clusters in 93.1% in the Bangladesh subregion, 85.7% in Cambodia, and 71.1% in the Kenya subregion across evaluated combinations, due to pathogen prevalences being omitted from the estimation of Q(s) when $d_{i,j} = 0$. The smallest differences between the two approaches were observed in Kenya, where several high-burden pathogens also share an exact intervention (e.g., malaria species, measles and rubella), and the largest differences were observed for combinations of pathogens with no shared intervention, such as interventions entailing tetanus immunization in Cambodia, whose burden does not contribute to Q(s) under the shared-intervention matrix.

Under the shared-intervention matrix, we identified the most efficient integrated interventions that included a vaccine-preventable disease in each setting. In the Bangladesh subregion, the same strategy was identified to be most efficient (measles and rubella vaccination campaign integrated with mass drug administration for soil-transmitted helminths), likely due to many shared interventions across intervenable pathogens. In the Kenya subregion, the most efficient strategy combined measles vaccination with malaria control (*Plasmodium falciparum, Plasmodium malariae, and Plasmodium ovale*), and in Cambodia, the identified strategy integrated a tetanus booster campaign with malaria control (*Plasmodium falciparum and Plasmodium vivax).* The figure below compares efficiency, as measured by the minimum number of spatial clusters reached to target 80% of all intervention need, for integrated strategies using both matrices.

**Figure S5.** The proportion of spatial clusters to reach 80% of all intervention beneficiaries when $d_{i,j}$ =1 for all pathogen pairs, in line with the main text analysis (x-axis), compared to using the shared-intervention $d_{i,j}$ (y-axis), where each individual point corresponds to an integrated combination, with points above the y = x line indicating more efficient targeting under the approach used in the main analysis. Integrated interventions falling close to the y = x line represent integration strategies in which prioritizing clusters by shared interventions reached 80% of burden nearly as efficiently as prioritizing by only spatial co-occurrence, indicating potential operational feasibility in addition to spatial feasibility.

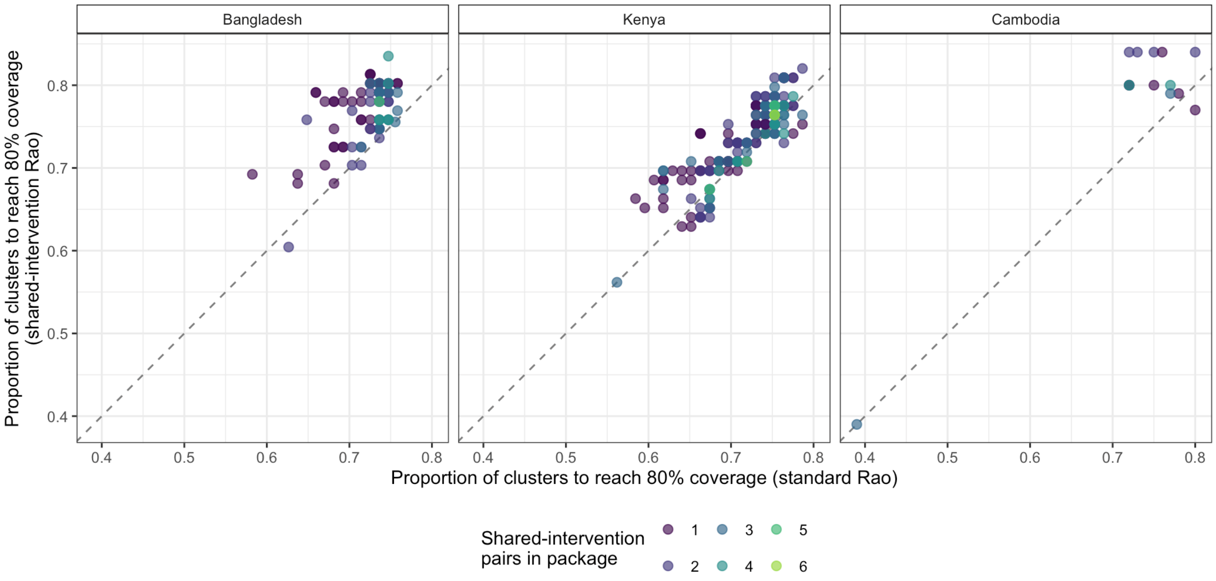

**Supplementary Methods 2. Multidimensional poverty index calculation.** Multidimensional poverty was calculated using the methodology established by the United Nations Development Programme[9,10]. The measure is estimated across ten core indicators *i* across three strata: health, education and standards of living, which are measured binarily (g = [0,1]) and assigned a specified weight *w*. $\sum w_i\ g_i$ was calculated for each individual/household unit from population metadata in each location. We calculated *H,* the proportion of households/individuals who were above an established threshold indicating that the households in a given spatial cluster were deprived in at least a third of weighted indicators. The cluster-level intensity of poverty A was calculated by averaging $\sum w_i\ g_i$ across all units above this threshold. Finally, cluster-level multidimensional poverty is set equal to A x H (proportion of impoverished x intensity of impoverishment among those defined as being impoverished).

**Table S3. Variables used to calculate multidimensional poverty by setting.**

| Indicator | Definition | Weight | Bangladesh | Kenya | Cambodia |
|---|---|---|---|---|---|
| Nutrition | Deprived if anyone in household is reported as undernourished or food insecure | 1/6 | Hficat: food security | HHS: hunger score<br>HHS_bi: hunger dichotomized | ha11: weight for height standard deviation<br><br>Ha40: body mass index |
| Child mortality | Any child has died in the household in the last five years preceding the study | 1/6 | NA | Isdead_el: child died before endline<br><br>Childdeath: child death after delivery | Hv111: mother alive<br>Hv113: father alive |
| Education | A household member has not completed at least six years of schooling, school-age children not attending school | 1/6 | Momedu: mom education level<br><br>Momeduy: education in years<br><br>Dadeduy: education in years | Motheredu: maternal education category<br><br>Fahteredu: Paternal education categories | Hv106: highest education level<br>Hv107: highest year of education<br>Hv109: educational attainment<br>Ha67: womans highest year of education |
| Cooking fuel | Household cooks with dung, | 1/18 | NA | NA | Hv226: type |

| | | | | | |
|---|---|---|---|---|---|
| | wood, charcoal, or coal | | | | of cooking fuel |
| Sanitation | Household sanitation facility is not improved, or improved but shared with other households (Sustainable Development Goals- SDG) | 1/18 | latown: owns a latrine<br>Latfeces: feces on latrine<br>Latseal: latrine has seal<br><br>OR treatment = sanitation | Imp_lat_bl: household access to improved latrine<br><br>OR treatment = sanitation | Hv205: type of toilet facility<br>Hv225: share toilet with other households |
| Water | No access to improved water source or within 30 min of water source (SDG) | 1/18 | Watmin: distance to water<br>Tubewel: improved water source<br>Storewat: water stored safely<br><br>OR treatment = water | Water_time: time to primary water source<br><br>Prim_drink_ws_bl: water source improved<br><br>OR treatment = water | Hv237: anything done to make water safe to drink<br>Sh104: time to get to water source in dry season<br>Sh104b: time to get to water source in wet season |
| Electricity | The household has no electricity | 1/18 | elec | elec | Hv206: has electricity |
| Housing | At least one of three housing materials for roof, walls, and floor are inadequate (of natural materials) | 1/18 | Floor<br>Walls<br>Roof | Floor<br>Walls<br>Roof | Hv213: main floor material<br>Hv214: main wall material<br>Hv215: main roof material |
| Assets | The household does not own more than one of these assets: radio, television, telephone, computer, animal cart, bicycle, motorbike/refrigerator, car/truck | 1/18 | Tv, refrigerator, bike, motor, phone | Tv, mobilephone, bicycle, car | Hv208: tv, Hv209: fridge<br>Hv210: bicycle, hv211: motorcycle, Hv212: car, Hv221: has a landline phone, Hv243c: animal drawn cart |
